# Differential Non-Coding RNA Profiles for Lung Cancer Early Detection in African and White Americans

**DOI:** 10.1101/2024.03.27.24304977

**Authors:** Lu Gao, Pushpa Dhilipkannah, Van K Holden, Janaki Deepak, Ashutosh Sachdeva, Nevins W Todd, Sanford A Stass, Feng Jiang

## Abstract

**Introduction:** Lung cancer leads in cancer-related deaths. Disparities are observed in lung cancer rates, with African Americans (AAs) experiencing disproportionately higher incidence and mortality compared to other ethnic groups. Non-coding RNAs (ncRNAs) play crucial roles in lung tumorigenesis. Our objective was to identify ncRNA biomarkers associated with the racial disparity in lung cancer.

**Methods:** Using droplet digital PCR, we examined 93 lung-cancer-associated ncRNAs in the plasma and sputum samples from AA and White American (WA) participants, which included 118 patients and 92 cancer-free smokers. Subsequently, we validated our results with a separate cohort comprising 56 cases and 72 controls.

**Results:** In the AA population, plasma showed differential expression of ten ncRNAs, while sputum revealed four ncRNAs when comparing lung cancer patients to the control group. In the WA population, the plasma displayed eleven ncRNAs, and the sputum had five ncRNAs showing differential expression between the lung cancer patients and the control group. For AAs, we identified a three-ncRNA panel (plasma miRs-147b, 324-3p, 422a) diagnosing lung cancer in AAs with 86% sensitivity and 89% specificity. For WAs, a four-ncRNA panel was developed, comprising sputum miR-34a-5p and plasma miRs-103-3p, 126-3p, 205-5p, achieving 88% sensitivity and 87% specificity. These panels remained effective across different stages and histological types of lung tumors and were validated in the independent cohort.

**Conclusions:** The ethnicity-related ncRNA signatures have promise as biomarkers to address the racial disparity in lung cancer.

## Introduction

Lung cancer is the leading cause of cancer-related deaths in both men and women in the USA ^1^. Non-small cell lung cancer (NSCLC) accounts for 85% of all lung cancer cases and is mainly composed of two histological types: adenocarcinoma (AC) and squamous cell carcinoma (SCC) ^1^. Early detection and timely treatment can significantly reduce morbidity and mortality in NSCLC ^1^. However, existing diagnostic methods fall short in the early detection of NSCLC. Furthermore, there are notable disparities in NSCLC between different ethnicities, with African Americans (AAs) experiencing a higher prevalence and mortality rate from the disease ^2^. The annual incidence of lung cancer is highest among AAs at 76.1 per 100,000, followed by White Americans (WAs) at 69.7 per 100,000, American Indians/Alaska Natives at 48.4 per 100,000, and Asian/Pacific Islanders at 38.4 per 100,000 ^2^. In addition to socioeconomic differences, biological factors like tumor biology, genetics, and molecular alterations also contribute to the disparities in lung cancer ^3^. For instance, genome-wide association studies have identified lung cancer susceptibility loci on chromosomes 5p15 and 15q25 in an AA population^4^.

Differences in the methylation levels of genes with functional relevance, like the nuclear receptor subfamily 3, have been identified as potential contributors to racial disparities in NSCLC ^5^. The epidermal growth factor receptor (EGFR) mutation is more common in AA lung cancer patients compared to other populations^6^. mRNA transcripts from AAs are less likely to undergo alternative polyadenylation in lung cancer compared to WAs ^7^. Furthermore, elevated levels of cytokines, including IL-1β, IL-10, and TNFα, are associated with an increased risk of lung cancer in the AA population ^8^. The molecular and genetic variations linked to lung cancer disparities offer potential as biomarkers for NSCLC in AAs, which could address the observed ethnic disparities in lung cancer treatment and outcomes ^9^.

Non-coding RNAs (ncRNAs) are RNA molecules that are not translated into proteins but are essential in regulating gene expression and cellular processes ^10^. ncRNAs mainly consist of microRNAs (miRNAs), long non-coding RNAs (lncRNAs), and small nucleolar RNAs (snoRNAs), among others^11^. Aberrant expression of certain ncRNAs has been closely linked to the onset and advancement of cancer, highlighting their potential as both therapeutic targets and diagnostic markers. Furthermore, miRNAs are implicated in the disparities observed between AAs and other populations^10, 11^. Numerous studies, ours included, have identified miRNAs, lncRNAs, and snoRNAs linked to lung cancer, underscoring their promise as potential biomarkers for this disease ^12-37^. We postulate that by studying and distinguishing ncRNA patterns in the plasma and sputum of NSCLC patients from both AA and WA cohorts, we can craft noninvasive lung cancer biomarkers tailored to individual ethnicities.

## MATERIAL and METHODS

### Patients and research design

The study received approval from the Institutional Review Board of the University of Maryland Baltimore. Participants eligible for inclusion were current or former smokers aged between 50 and 80 years old. We excluded those who were pregnant or lactating, those with current pulmonary infections, individuals who had undergone thoracic surgery within the last 6 months, those who had received chest radiotherapy in the preceding year, and patients with a life expectancy of under one year. Demographic and clinical data, including age, sex, race, and smoking history, were collated from medical records. To confirm malignancy, tissue samples obtained either surgically or via biopsy were subject to pathologic examination. Surgical pathologic staging adhered to the TNM classification of the International Union Against Cancer, the American Joint Committee on Cancer, and the International Staging System for Lung Cancer. Histopathologic classification was made based on the guidelines provided by the World Health Organization. Radiographic characteristics of the pulmonary nodules (PNs) were derived from CT images. These included the maximum transverse size, the visually determined type (categorized as nonsolid, ground-glass opacity, part-solid, solid, peri fissure, or spiculation), and the nodule’s lung location. A benign diagnosis was confirmed either pathologically, specifying a benign cause, or through the clinical and radiographic stability of the PNs observed across multiple check-ups over a 2-year follow-up period. 340 participants were enrolled. Among the 340 participants, 174 patients were diagnosed with NSCLC, with an equal distribution of 87 AAs and 87 WA lung cancer patients. The other 166 had benign conditions: 99 had granulomatous inflammation, 38 exhibited nonspecific inflammatory changes, and 29 presented with lung infections. The cohort was bifurcated randomly into a training set and a validation set, detailed in Tables 1 and 2.

**Table 1.** Characteristics of a training set of NSCLC patients and cancer-free smokers.

|  |  |  |
| --- | --- | --- |
|  | NSCLC cases (n = 118) | Controls (n = 92) |
| Age | 67.25 (SD 12.33) | 66.29 (SD 11.19) |
| Sex |  |  |
| Female | 46 | 36 |
| Male | 72 | 56 |
| Race |  |  |
| African Americans | 59 | 46 |
| White Americans | 59 | 46 |
| Smoking pack-years (median) | 31.6 | 30.9 |
| Pulmonary nodule size (mm) | 19.37 (SD 12.16) | 6.12 (SD 4.58) |
| Stage |  |  |
| Stage I | 78 |  |
| Stage II | 26 |  |
| Stage III-IV | 14 |  |
| Histological type |  |  |
| Adenocarcinoma | 69 |  |
| Squamous cell carcinoma | 49 |  |
| Abbreviations: NSCLC, non-small cell lung cancer. SD, standard deviation. |  |  |

**Table 2.** Characteristics of a validation set of NSCLC patients and cancer-free smokers.

|  |  |  |
| --- | --- | --- |
|  | NSCLC cases (n = 56) | Controls (n = 72) |
| Age | 67.84 (SD 11.73) | 67.91 (SD 11.07) |
| Sex |  |  |
| Female | 20 | 25 |
| Male | 36 | 47 |
| Race |  |  |
| African Americans | 28 | 36 |
| White Americans | 28 | 36 |
| Smoking pack-years (median) | 35.1 | 34.6 |
| Pulmonary nodule size (mm) | 20.27 (SD 11.46) | 7.48 (SD 5.13) |
| Stage |  |  |
| Stage I | 26 |  |
| Stage II | 24 |  |
| Stage III-IV | 6 |  |
| Histological type |  |  |
| Adenocarcinoma | 32 |  |
| Squamous cell carcinoma | 24 |  |
| Abbreviations: NSCLC, non-small cell lung cancer. SD, standard deviation. |  |  |

### Sputum and blood sample collection and preparation

Specimens were obtained from participants prior to any treatment initiation. For sputum collection, participants were instructed to blow their nose, rinse their mouth, and drink water to minimize contamination from oral epithelial cells. The sputum was then gathered in sterile containers and promptly processed on ice using 0.1% dithiothreitol and phosphate-buffered saline (Sigma Aldrich, St. Louis, MO) ^12-33^. Concurrently, blood samples were drawn, and within an hour of collection, plasma was separated following standard clinical protocols ^12-33^.

### RNA isolation

RNA was extracted from the specimens using the miRNeasy Mini Kit spin column (QIAGEN, Germantown, MD), as previously described ^12-33^. The extracted RNA samples were promptly stored at -80°C in barcoded cryotubes.

### Droplet digital PCR (ddPCR) analysis of miRNAs, lncRNAs, and snoRNAs

Numerous studies, including our own, have pinpointed 93 specific miRNAs, lncRNAs, and snoRNAs in tissue specimens related to lung cancer, suggesting their potential as valuable biomarkers for the disease ^12-37^ (Supplementary table 1). In this study, we employed ddPCR analysis for the 93 ncRNAs in both plasma and sputum, following the previously developed methods ^12-33^. One μl of RNA from each sample was reverse transcribed (RT) using gene-specific primers for each target with the TaqMan miRNA RT Kit (Applied Biosystems, Foster City, CA). For the ddPCR reactions, a mixture containing 5 μl cDNA solution, 10 μl Supermix, and 1 μl Taqman primer/probe mix was prepared in a 20 μl volume. This mixture was loaded into cartridges filled with droplet generation oil (Bio-Rad, Hercules, CA) and placed into the QX100 Droplet Generator (Bio-Rad). The droplets formed were then shifted to a 96-well PCR plate, followed by PCR amplification using a T100 thermal cycler (Bio-Rad). The ddPCR method generated over 10,000 droplets per well, which were subsequently analyzed using a fluorescence detector. This ensured that the ncRNAs were consistently and accurately detected in the clinical samples. We assessed the number of positive reactions and employed Poisson’s distribution to accurately determine the concentration of the target genes^12-33^.

### Statistical analysis

Statistical significance for biomarkers and clinical determinants was ascertained using the Mann-Whitney U test or the Chi-Square test. Pearson’s correlation was used to assess the relationship between miRNA expression and clinical and demographic data, including smoking history measured as pack-years. The construction of a lung cancer biomarker panel was strategized by bifurcating the cohort into training and validation subsets, adhering to the guidelines proposed by the National Cancer Institute’s Early Detection Research Network. In the training subset, feature selection was conducted employing the LASSO (Least Absolute Shrinkage and Selection Operator) in tandem with logistic regression. A 10-fold cross-validation reinforced with bootstrapping was used to mitigate outlier influence. The mean decrease in Gini impurity served as the metric for evaluating variable importance, with the False Discovery Rate (FDR) addressing multiple testing corrections. Discrimination metrics were established using the ROC (receiver operating characteristic) curve analysis, reporting AUC (area under the curve) values accompanied by 95% confidence intervals. The confidence intervals for performance metrics (AUC, sensitivity, and specificity) were determined employing an assortment of statistical methodologies. After refining the diagnostic panels from the training set, we assessed their robustness in the validation subset using AUC, sensitivity, and specificity.

## RESULTS

### Differential expression of ncRNAs in plasma and sputum of NSCLC patients vs. cancer-free smokers

The expression levels of 93 lung cancer-associated ncRNAs^12-37^, which included 67 miRNAs, 21 snoRNAs, and five lncRNAs (Supplemental Table 1), were quantified in plasma and sputum using a microplate-based ddPCR technique^12-35^. This analysis was first conducted on the specimens obtained from 59 AA NSCLC patients, 46 cancer-free AA smokers, 59 WA NSCLC patients, and 46 cancer-free WA smokers (Table 1). In plasma, a differential expression of 25 ncRNAs, including 18 miRNAs, five snoRNAs and two lncRNAs, was observed between cancer patients and cancer-free smokers across all ethnicities (Mann-Whitney U test: p < 0.05; FDR-adjusted p < 0.05) (Supplemental Table 2). Similarly, in sputum, eight ncRNAs – comprising three miRNAs, three snoRNAs, and two lncRNAs – exhibited differential expression between NSCLC patients and cancer-free smokers (Mann-Whitney U test: p < 0.05; FDR-adjusted p < 0.05) (Supplemental Table 2).

### Differential expression of ncRNAs in plasma and sputum among individuals from different ethnic populations

We further explored the differential expression of ncRNAs in plasma and sputum across individual ethnic groups. In plasma samples from AA participants, seven ncRNAs (miRs-31-5p, 147b, 16-5p, 375-3p, 422a, and 324-3p, and snoRA42) showed a significant increase in expression in lung cancer patients compared to cancer-free AA controls (Mann-Whitney U test: p < 0.05; FDR-adjusted p < 0.05) (Table 3, Figure 1, and Supplemental Figure 1). However, snoRA76 displayed a significant decrease in expression in AA lung cancer patients compared to controls (Mann-Whitney U test: p < 0.05; FDR-adjusted p < 0.05) (Table 3, Figure 1, and Supplemental Figure 1). In the sputum samples from AA participants, four ncRNAs – three microRNAs (miRs-16-5p, 210-3p, and 205-5p) and one snoRNA (snoRA116) – were found to be elevated in lung cancer patients as compared to cancer-free AA controls (Mann-Whitney U test: p < 0.05; FDR-adjusted p < 0.05) (Table 3 and Figure 1A) (Supplemental Figure 1).

**Table 3.** The set of 12 ncRNAs found to be significantly differential expressed in AAs, as analyzed by the Mann-Whitney U test, with a False Discovery Rate (FDR)-adjusted p-value of less than 0.05.

| ncRNAs | Mean Expression (Cancer Patients) | Mean Expression (Controls) | Mann-Whitney U Statistic | FDR-adjusted p-value |
| --- | --- | --- | --- | --- |
| Plasma miR-31-5p | 2.115 | 1.8130 | 864 | 0.0005 |
| Plasma miR-147b | 1.700 | 1.3100 | 875 | 0.0016 |
| Plasma miR-16-5p | 2.793 | 1.7890 | 953 | 0.0086 |
| Plasma miR-375-3p | 2.413 | 1.7600 | 870 | 0.0015 |
| Plasma miR-422a | 2.376 | 1.9640 | 827 | 0.0005 |
| Plasma miR-324-3p | 2.470 | 2.0100 | 1232 | 0.0126 |
| Plasma snoRA42 | 1.783 | 1.5700 | 1336 | 0.0407 |
| Plasma snoRA76 | 1.391 | 2.1990 | 1600 | 0.0205* |
| Sputum miR-210-3p | 2.461 | 1.6780 | 1485 | 0.0041 |
| Sputum snoRA116 | 2.315 | 1.7880 | 1480 | 0.0038 |
| Sputum miR-16-5p | 2.793 | 1.7660 | 1374 | 0.0007 |
| Sputum miR-205-5p | 2.647 | 1.9840 | 1284 | 0.0001 |

\*, the ncRNA was significantly decreased in expression in AA lung cancer patients compared to cancer-free AA smokers.

**Figure 1.**
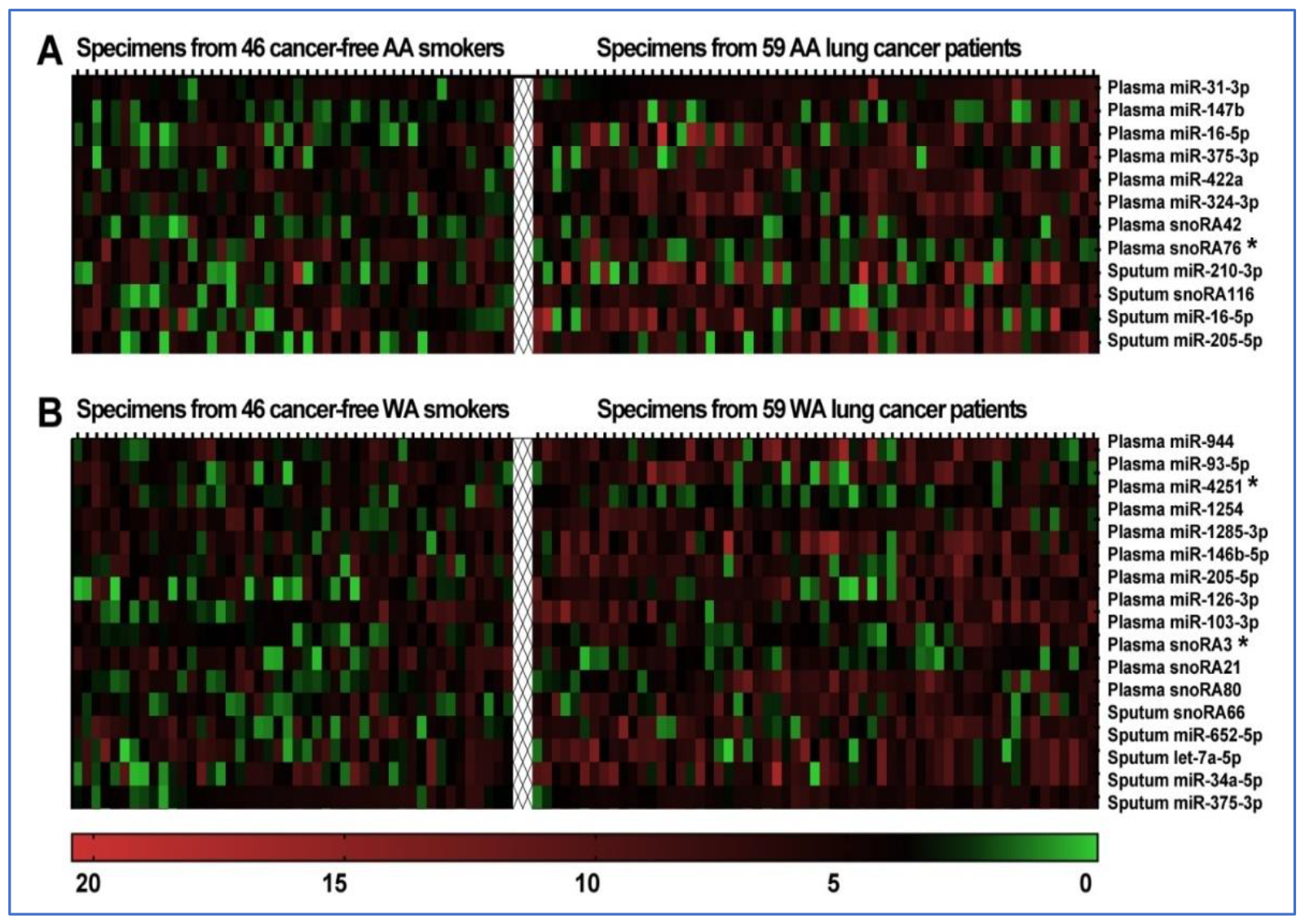
Differential expression heatmap of ncRNAs in plasma and sputum samples from AA and WA lung cancer patients versus their healthy counterparts. **A**. Among 59 AA lung cancer patients and 46 cancer-free AA smokers, 12 ncRNAs showed statistically significant expression differences (all p <0.05). **B**. In a comparison between 59 WA lung cancer patients and 46 cancer-free WA smokers, 17 genes revealed statistically significant expression differences with a p-value <0.05. The color scale spans from green (indicating down-regulation) to black (no change) and red (signifying up-regulation). * ncRNAs with statistically significant reductions in expression in lung cancer patient specimens compared to specimens from cancer-free smokers.

In plasma samples from WA participants, 12 ncRNAs showed differential expression between lung cancer patients and cancer-free WA smokers. These ncRNAs include eight microRNAs (miRs-93-5p, 103a-3p, 126-3p, 146b-5p, 205-5p, 944, 4251, and 1285-3p) and three snoRNAs (snoRA3, snoRA21, and snoRA80) (Mann-Whitney U test: p < 0.05; FDR-adjusted p < 0.05) (Table 4 and Figure 1B) (Supplemental Figure 2). Ten of these ncRNAs (miRs-93-5p, 103a-3p, 126-3p, 146b-5p, 205-5p, 944, 1254, and 1285-3p, and snoRA21 and snoRA80) showed increased expression, whereas two ncRNAs (miR-4251 and snoRA3) had decreased expression in WA lung cancer patients when compared to cancer-free AA smokers. In sputum samples from WAs, five ncRNAs demonstrated elevated expression in lung cancer patients compared to cancer-free WA smokers. These ncRNAs include four miRNAs: miR-34a-5p, miR-652-5p, miR-375-3p, and let-7a, along with one snoRNA: snoRA66 (Mann-Whitney U test: p < 0.05; FDR-adjusted p < 0.05) (Table 4, Figure 1, and Supplemental Figure 2).

**Table 4.** The set of 17 ncRNAs found to be significantly differential expressed in WAs, as analyzed by the Mann-Whitney U test, with a False Discovery Rate (FDR)-adjusted p-value of less than 0.05.

| ncRNAs | Mean Expression (Cancer Patients) | Mean Expression (Controls) | Mann-Whitney U Statistic | FDR-adjusted p-value |
| --- | --- | --- | --- | --- |
| Plasma miR-944 | 2.279 | 1.738 | 897 | 0.0027 |
| Plasma miR-93-5p | 2.325 | 1.531 | 1007 | 0.0123 |
| Plasma miR-4251 | 1.328 | 1.826 | 1003 | 0.0115* |
| Plasma miR-1254 | 2.162 | 1.595 | 1032 | 0.0262 |
| Plasma miR-1285-3p | 2.531 | 1.947 | 884 | 0.002 |
| Plasma miR-146b-5p | 2.529 | 1.628 | 975 | 0.0132 |
| Plasma miR-205-5p | 2.228 | 1.489 | 911 | 0.0037 |
| Plasma miR-126-3p | 2.333 | 1.714 | 773 | 0.0001 |
| Plasma miR-103-3p | 1.681 | 1.315 | 909 | 0.0051 |
| Plasma snoRA3 | 1.195 | 1.473 | 932 | 0.0057* |
| Plasma snoRA21 | 2.317 | 1.618 | 875 | 0.0017 |
| Plasma snoRA80 | 2.226 | 1.546 | 846 | 0.0008 |
| Sputum snoRA66 | 2.385 | 1.881 | 1038 | 0.0213 |
| Sputum miR-652-5p | 2.463 | 1.984 | 1027 | 0.0325 |
| Sputum let-7a-5p | 2.376 | 1.555 | 907 | 0.0034 |
| Sputum miR-34a-5p | 1.998 | 1.541 | 1024 | 0.0317 |
| Sputum miR-375-3p | 2.137 | 1.844 | 984 | 0.0157 |

\*, the ncRNA was significantly decreased in expression in AA lung cancer patients compared to cancer-free AA smokers.

**Figure 2.**
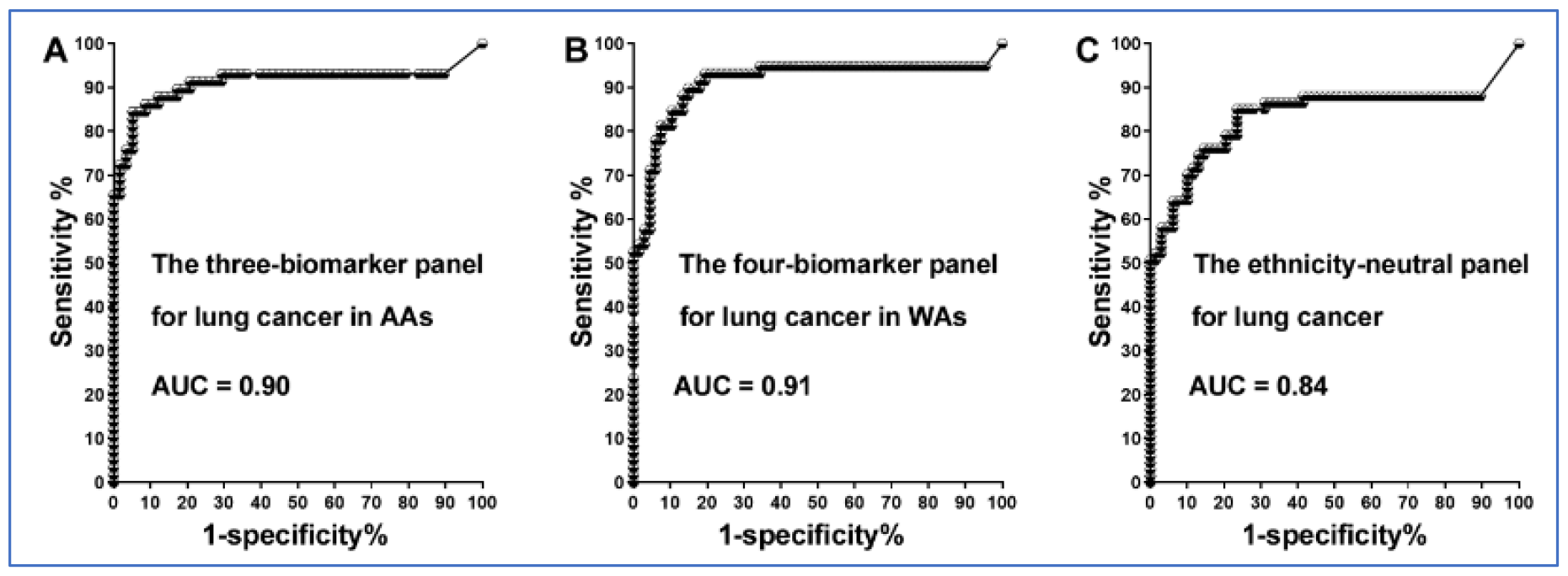
Performance of the three individual panels for diagnosing lung cancer in different ethnic groups in the training set. **A**. ROC curves depict the accuracy of the three-biomarker panel in diagnosing lung cancer in AAs, yielding an AUC of 0.90. **B**. ROC curves represent the accuracy of the four-biomarker panel in diagnosing lung cancer in WAs, with an AUC of 0.91. **C**. The pan-ethnic biomarker panel displays an AUC of 0.84 in the diagnosis of lung cancer, irrespective of racial populations.

### The diagnostic utility of the plasma and sputum ncRNA biomarkers vary with ethnicity

We utilized logistic regression and a backward elimination approach to identify specific ncRNA biomarker panels for lung cancer in different ethnicities. For AAs, the best prediction for lung cancer was achieved using a combination of three ncRNAs: miRs-147b, 324-3p, and 422a in plasma. This panel yielded an AUC of 0.90, distinguishing AA cancer patients from healthy AAs with a sensitivity of 86% and a specificity of 89% (Fig. 2) (Table 5). For WAs, the optimal prediction derived from a combination of four ncRNAs: sputum miR-34a-5p, plasma miR-103-3p, plasma 126-3p, and plasma 205-5p. This combination achieved an AUC of 0.91, diagnosing NSCLC with a sensitivity of 89% and a specificity of 87% (all p < 0.05) (Table 5). Additionally, for pan-ethnic diagnosis, a panel consisting of plasma miR-21-3p, plasma miR-210-3p, and sputum miR-126-3p demonstrated the best universal diagnostic ability. This combination achieved an AUC of 0.84, with a sensitivity of 71% and a specificity of 88% (Fig. 2) (Table 5). The pan-ethnic biomarker panel exhibited lower sensitivity for AAs and WAs compared to their individual biomarker panels (71% vs. 86% for AAs and 89% for WAs, p<0.05), while maintaining similar specificity (Table 5). Among the ten ncRNA biomarkers, plasma miR-205-5p and sputum miR-126-3p were associated with age, whereas plasma miR-422a and plasma miR-324-3p were related to the patients’ sex (all p-values < 0.05) (Supplement Table 3). Plasma miR-422a was associated with the size of PNs, and plasma miR-147b correlated with tumor stage. The ncRNAs were not linked to smoking history (Supplement Table 3). When these biomarkers were used in combination as panels, their diagnostic values did not show any association with the patients’ age, sex, smoking history, size of PNs, tumor stages, or histological types of lung tumors.

**Table 5.** The diagnostic values of the three individual panels in the training set.

| Diagnostic performance of the three-biomarker panel in AAs |  | Diagnostic performance of the four-biomarker panel in WAs |  | Diagnostic performance of the pan-ethnic biomarker panel |  |
| --- | --- | --- | --- | --- | --- |
| Sensitivity, % (95% CI) | Specificity, % (95% CI) | Sensitivity, % (95% CI) | Specificity, % (95% CI) | Sensitivity, % (95% CI) | Specificity, % (95% CI) |
| 86.44% (75.02% to 93.96%) | 89.13% (76.43% to 96.38%) | 89.83% (79.17% to 96.18%) | 86.96% (73.74% to 95.06%) | 71.19% (62.13% to 79.15%) | 88.04% (79.61% to 93.88%) |

### Verifying the diagnostic potential of the biomarker panels for disparities

We validated the three distinct ncRNA biomarker panels for diagnosis of lung cancer in the validation cohort. For AAs, the biomarker panel comprising plasma miR-147b, miR-324-3p, and miR-422a achieved a sensitivity of 86% and a specificity of 89% in detecting lung cancer (Supplementary Table 4). For WAs, the panel including sputum miR-34a-5p, plasma miR-103-3p, plasma 126-3p, and plasma 205-5p demonstrated a sensitivity of 89% and a specificity of 86% (Supplementary Table 4). The ethnicity-neutral biomarker panel, including plasma miR-21-3p, plasma miR-210-3p, and sputum miR-126-5p, achieved a sensitivity of 71% and 89% specificity in the diagnosis of lung cancer across all ethnic groups (Supplementary Table 4). These results in the validation set confirm the findings in the training set and thus support the potential of these biomarkers for early NSCLC detection in different racial populations.

## DISCUSSION

The National Lung Screening Trial (NLST) has established that low-dose computed tomography (LDCT) screenings significantly reduce lung cancer-related mortality among high-risk populations, notably smokers ^38^. LDCT is currently utilized for lung cancer screening in smokers. However, this method has significantly increased the detection of indeterminate pulmonary nodules (PNs) in asymptomatic individuals. Of the smokers screened, 24.2% were found to have indeterminate PNs through LDCT, yet 96.4% of these nodules were subsequently confirmed as benign growths^38^. Moreover, while the CT screening using LDCT boasts a sensitivity exceeding 90%, its specificity stands at a mere 61%(1), resulting in a substantial false positive rate or overdiagnosis^38^. Given the notably high incidence and mortality rates among AAs, there is an urgent need for non-invasive molecular biomarkers tailored for the AA demographic. These biomarkers can facilitate early detection of NSCLC either when used independently or in conjunction with LDCT, aiming to reduce the false positive rates frequently associated with LDCT. While the prior investigations have revealed certain miRNA variations in surgically resected lung tumor tissues between AA and WA patients ^11, 39^, the field still lacks non-invasive molecular biomarkers tailored for early lung cancer detection in the AA demographic.

In this study, we systematically analyzed 93 lung cancer-related ncRNAs from plasma and sputum samples of both AA and WA lung cancer patients, as well as from cancer-free controls. Distinct ncRNA alterations associated with each population were identified, leading to the formulation of specific diagnostic panels for each group. Moreover, we developed an ethnicity-neutral biomarker panel for diagnosing lung cancer. However, this pan-ethnic biomarker panel demonstrated suboptimal diagnostic sensitivity among varied ethnic groups compared to population-specific markers. Furthermore, while some ncRNAs are associated with age, gender, size of PNs, or smoking history, the combined use of these genes as biomarker panels was not influenced by these factors in either population. Interestingly, their diagnostic efficacy remained consistent across early and late stages of lung tumors, underscoring their potential for early NSCLC detection in clinical contexts. Additionally, these biomarkers are not associated with PNs identified via LDCT. Thus, these biomarkers may prove instrumental in distinguishing lung cancer within PNs identified by LDCT, potentially reducing its elevated false-positive rate. Nonetheless, a more extensive study with a broader cohort is essential to further validate this diagnostic potential.

The most discriminatory biomarkers that can diagnose lung cancer among AAs are miRs-147b, 324-3p, and 422a. miR-147b can promote lymph node metastasis and prognosis of cancer through its regulation of PRPF4B, WDR82, and NR3C2 ^40^. Additionally, it influences drug resistance to EGFR inhibitors by modulating the TCA cycle. miR-324-3p is highly present in lung cancer cells and promotes their growth and invasion ^41^. miR-422a can inhibit the TGF-β/SMAD pathway by downregulating sulfatase 2, and hence constrain NSCLC cell proliferation, migration, invasion, colony formation, EMT and tumorigenesis ^42^. miR-34a-5p, miR-103-3p, miR-126-3p, and 205-5p stand out in the diagnosis of lung cancer among WAs. miR-34-5p is implicated in the regulation of tumor growth due to its role in the epithelial-mesenchymal transition (EMT) via EMT-transcription factors, p53 and other important signal pathways^43^. Dysfunction of miR-103-3p is pivotal in lung tumorigenesis as it directly targets PDCD10, influencing lung cancer cell proliferation and metastasis^44^. The miR-103/PDCD10 signaling pathway offers a potential novel therapeutic target for NSCLC treatment^44^. miR-126-3p, an endothelial miRNA, is aberrantly expressed in specimens of patients with lung cancer ^45^. Its reintroduction curbs tumor growth by targeting EGFL7. Elevated expression of miR-205-5p is implicated in the initiation and progression of NSCLC ^46^.

This microRNA is also associated with the modulation of EMT by targeting EMT-related genes, which in turn affects the invasive and metastatic capabilities of lung cancer cells^47^. Furthermore, miR-205-5p is believed to contribute to the carcinogenesis and chemoresistance of NSCLC by influencing the PTEN signaling pathway^48^. Nevertheless, further investigation is needed to fully understand the specific roles and implications of these ncRNAs in accounting for racial disparities in lung cancer incidence.

This study might present valuable insights but also highlights areas that need further exploration. Sample sizes could certainly be expanded to uncover markers that are less discriminatory. Furthermore, while this study focused on analyzing 93 ncRNAs, a myriad of other genes awaits systematic validation in future work. In addition, a longitudinal study is warranted to investigate how these molecules relate to disease pathology and progression over time among the different race populations.

By analyzing surgically resected tissue samples, Mitchell et al. identified seven miRNAs with differing expression levels between AA and WA lung cancer patients ^39^. These miRNAs have limited similarity to the ones we identified using plasma and sputum samples. Several reasons could account for these discrepancies: the tissues provide localized information, whereas plasma and sputum reflect systemic influences. Inherent tumor variability can affect miRNA expression. Tumors may release specific ncRNAs based on their characteristics, with some remaining localized. Furthermore, variations in laboratory procedures and ncRNA detection methods might yield different results. In addition, genetic and environmental differences within AA and WA groups can influence ncRNA profiles across studies. In response to this discrepancy, we are currently collecting tissue specimens, matched with plasma and sputum samples, from various ethnic populations. This will enable us to concurrently profile ncRNA changes and better understand the relationship between molecular aberrations across these different specimen types.

In sum, the distinctive ncRNA profiles linked to lung cancer in AAs vs. WAs may hold promise as biomarkers to address the observed racial disparity in lung cancer. Nonetheless, a large multi-center clinical trial is needed to prospectively validate the full utility of the biomarkers for early lung cancer detection in the different populations.

## Data Availability

All data produced in the present study are available upon reasonable request to the authors

https://www.medschool.umaryland.edu/profiles/jiang-feng/

https://www.medschool.umaryland.edu/profiles/jiang-feng/

## Authorship Contribution Statement

**Lu Gao:** Conceptualization; Data curation; Investigation, Formal analysis; Methodology.

**Pushpa Dhilipkannah:** Data curation; Investigation, Formal analysis; Methodology.

**Van K Holden:** Data curation; Investigation, Formal analysis; Methodology. Writing—review and editing.

**Janaki Deepak:** Data curation; Investigation, Formal analysis; Methodology. Writing—review and editing.

**Ashutosh Sachdeva:** Data curation; Investigation, Formal analysis; Methodology. Writing—review and editing.

**Nevins W Todd:** Data curation; Investigation, Formal analysis; Methodology. Writing—review and editing.

**Sanford A Stass:** Data curation; Investigation, Formal analysis; Methodology. Writing—review and editing.

**Feng Jiang:** Conceptualization; Data curation; Formal analysis; Funding acquisition; Investigation; Methodology; Project administration; Resources; Supervision; Validation; Writing—original draft; Writing—review and editing.

## Ethics Approval and Consent to Participate

Ethical permits are in place for all studies and all participants consented to partake.

## Consent for Publication

All patients and authors consented to the publication of the results.

## Availability of Data and Materials

Data are available from the corresponding author on reasonable request.

## Acknowledgment

We extend our sincere gratitude to the Biostatistics Shared Service at the University of Maryland Marlene and Stewart Greenebaum Cancer Center for their invaluable contribution in conducting the statistical analysis for this study.

## Funding

Supported by NCI grant number: UH3 CA251139 (Dr. Feng Jiang).

## Disclosures

The authors declare no financial conflicts of interest or other competing interests related to this research.

## Supplemental files

**Supplemental Table 1.**
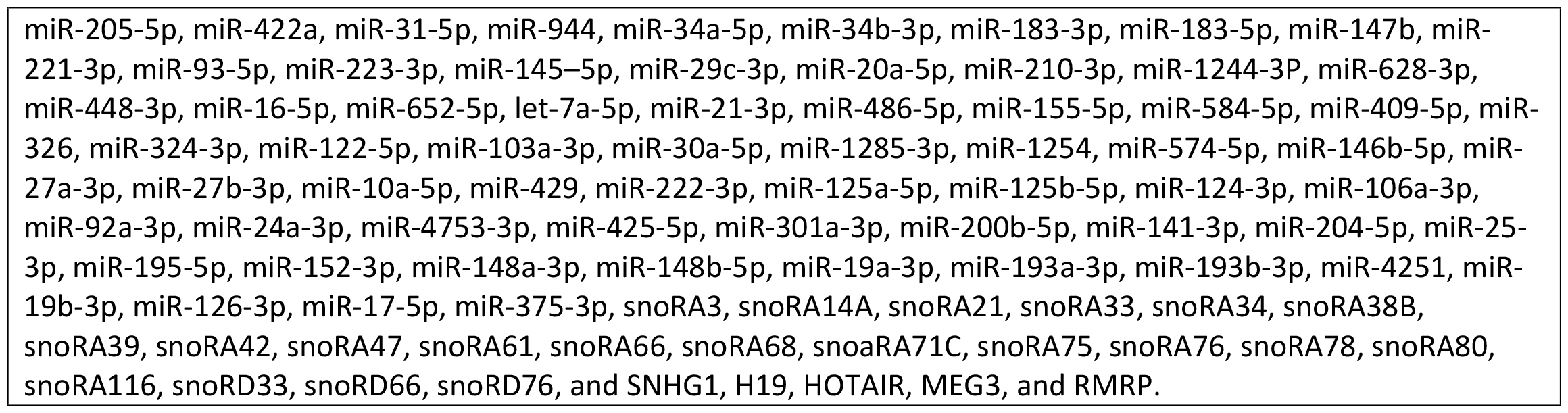
93 lung cancer-associated ncRNAs tested by ddPCR in this study.

**Supplemental Table 2.**
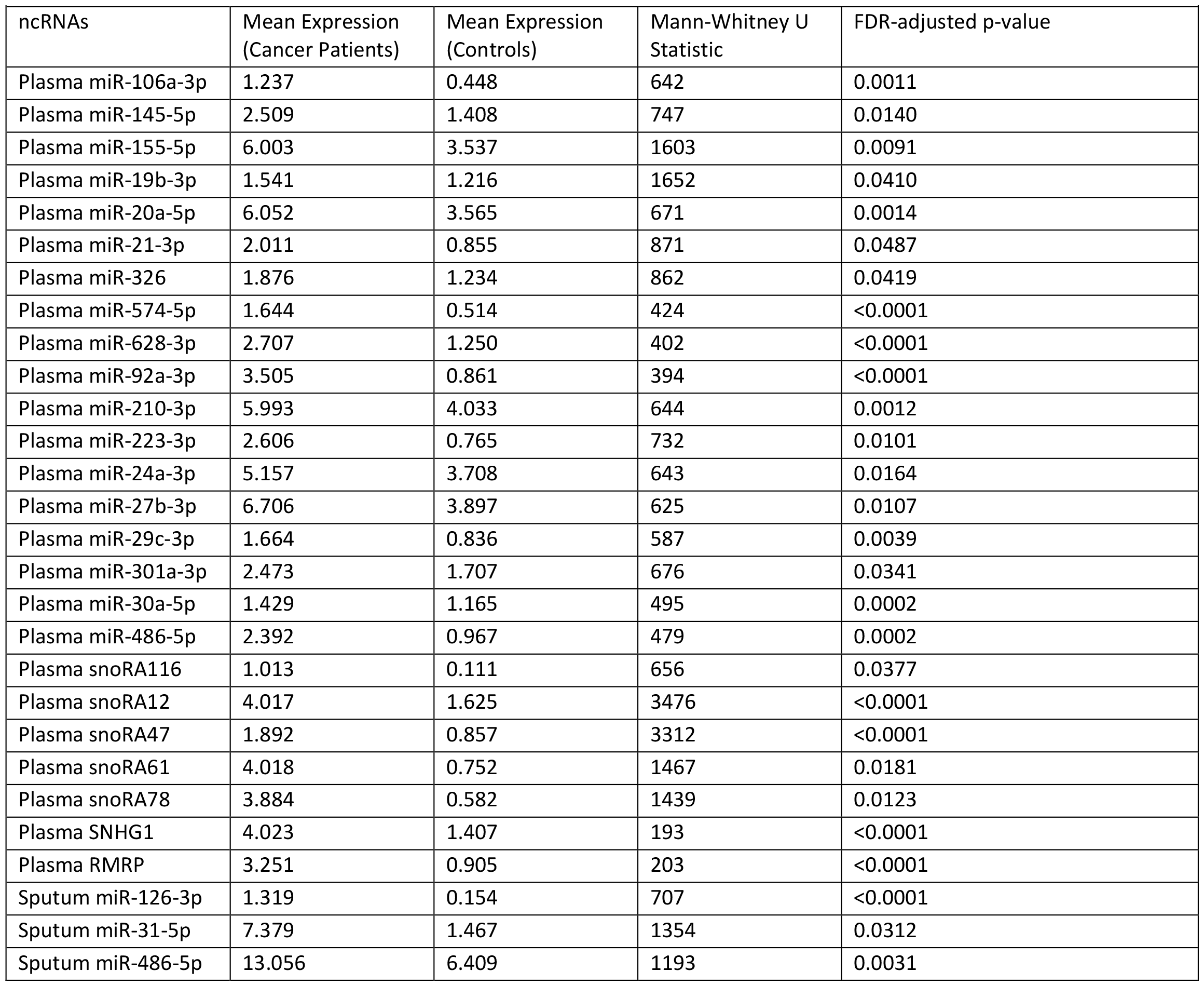
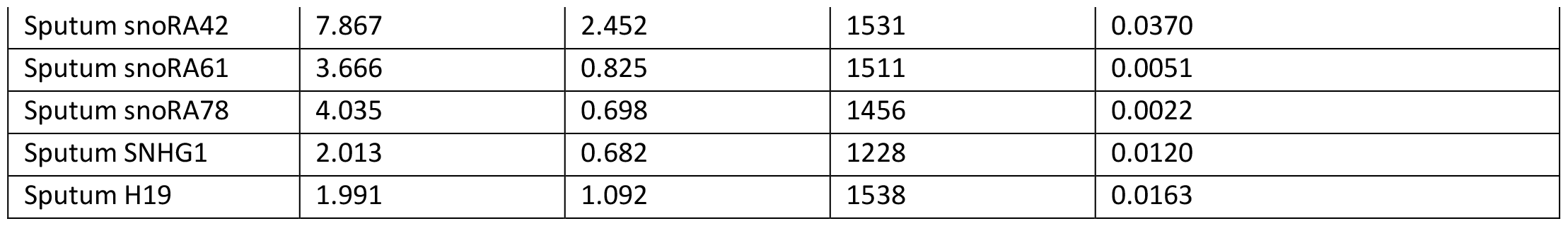
Among lung cancer patients and cancer-free smokers, 33 ncRNAs showed significantly differential expression in in plasma or sputum, irrespective of racial background, as analyzed by the Mann-Whitney U test, with a False Discovery Rate (FDR)-adjusted p-value of less than 0.05.

**Supplemental Table 3.** Associations between the ten ncRNAs in the biomarker panels and clinical and demographic data, analyzed using Pearson’s correlation coefficients.

| ncRNAs | Age | Sex | Smoking-Pack-Years | Pulmonary Nodule Size | Tumor Stage | Histology |
| --- | --- | --- | --- | --- | --- | --- |
| Plasma miR-422a | 0.738 | 0.016* | 0.250 | 0.018* | 0.562 | 0.456 |
| Plasma miR-324-3p | 0.995 | 0.001* | 0.588 | 0.060 | 0.752 | 0.264 |
| Plasma miR-147b | 0.472 | 0.414 | 0.877 | 0.091 | 0.008* | 0.819 |
| Plasma miR-205-5p | 0.048* | 0.555 | 0.596 | 0.834 | 0.835 | 0.779 |
| Plasma miR-126-3p | 0.186 | 0.624 | 0.408 | 0.130 | 0.905 | 0.555 |
| Plasma miR-103-3p | 0.571 | 0.191 | 0.983 | 0.055 | 0.934 | 0.561 |
| Sputum miR-34a-5p | 0.092 | 0.676 | 0.925 | 0.521 | 0.226 | 0.307 |
| Plasma miR-21-3p | 0.594 | 0.543 | 0.997 | 0.100 | 0.563 | 0.897 |
| plasma miR-210-3p | 0.200 | 0.115 | 0.614 | 0.542 | 0.313 | 0.688 |
| sputum miR-126-3p | 0.025* | 0.123 | 0.326 | 0.785 | 0.624 | 0.200 |

\* Significance at $p < 0.05$ .

**Supplemental Table 4.** The diagnostic values of the three individual panels in the validation set.

| Diagnostic performance of the three-biomarker panel in AAs |  | Diagnostic performance of the four-biomarker panel in WAs |  | Diagnostic performance of the pan-ethnic biomarker panel |  |
| --- | --- | --- | --- | --- | --- |
| Sensitivity, % (95% CI) | Specificity, % (95% CI) | Sensitivity, % (95% CI) | Specificity, % (95% CI) | Sensitivity, % (95% CI) | Specificity, % (95% CI) |
| 85.71% (67.33% to 95.97%) | 88.89% (73.94% to 96.89%) | 89.29% (71.77% to 97.73%) | 86.11% (70.50% to 95.33%) | 71.43% (57.79% to 82.70%) | 88.89% (79.28% to 95.08%) |

**Supplemental Figure 1.**
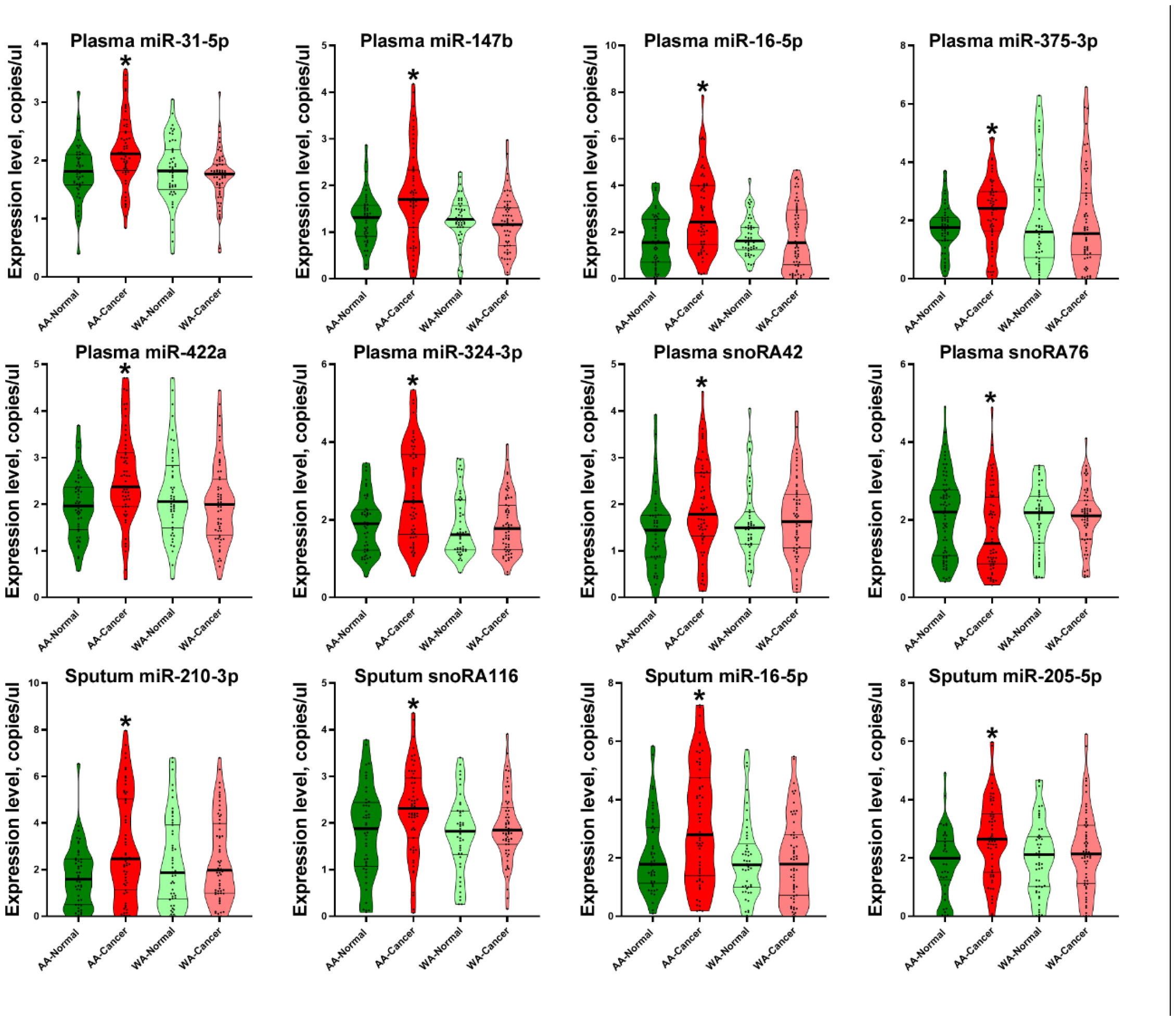
Differential expression of eleven ncRNAs in plasma and sputum between AA lung cancer patients and cancer-free smokers. *, p<0.05.

**Supplemental Figure 2.**
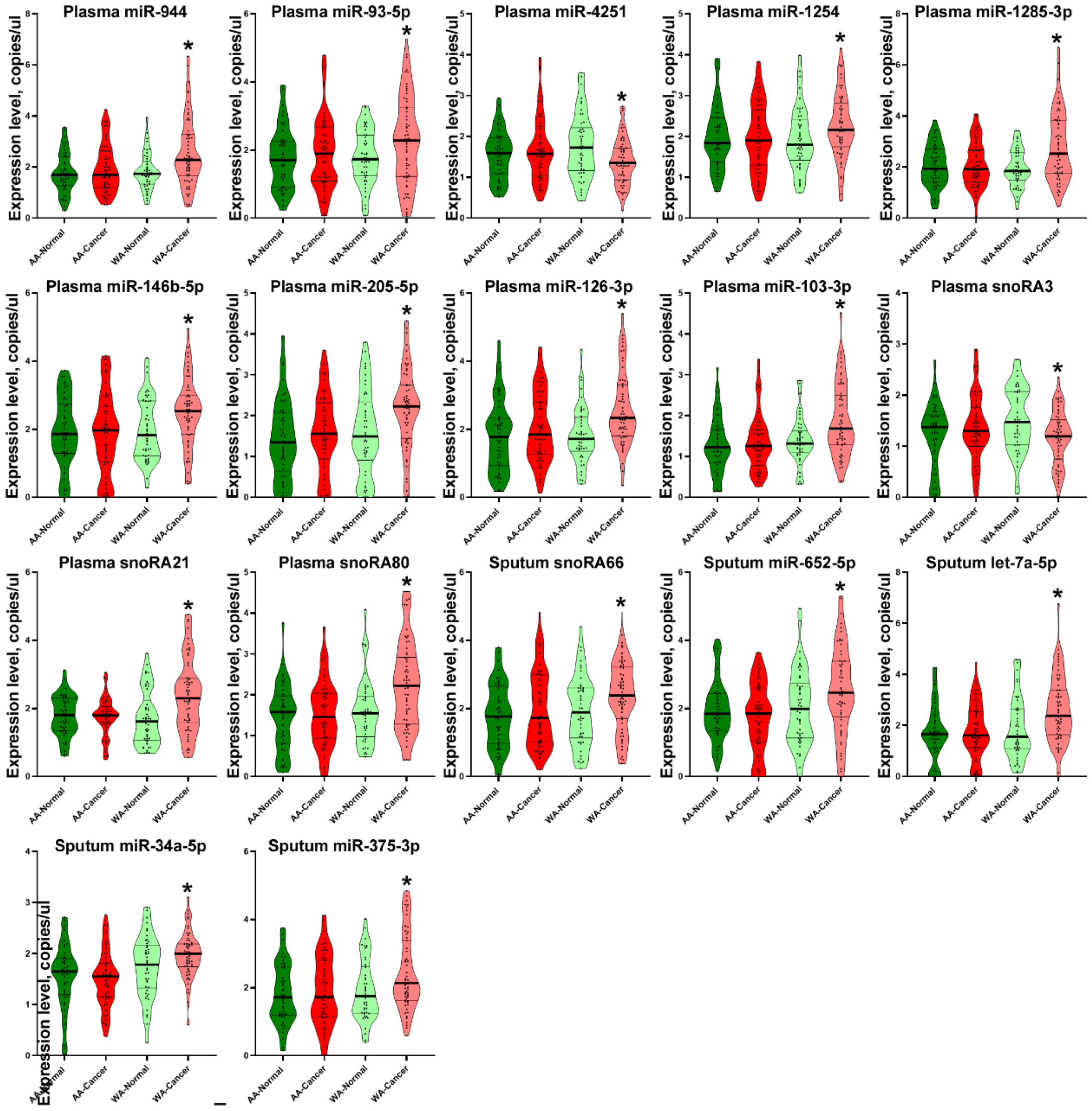
Differential expression of 16 ncRNAs in plasma and sputum between WA lung cancer patients and cancer-free smokers. *, p<0.05.

## Notes

### Competing Interest Statement

The authors have declared no competing interest.

### Clinical Protocols

https://www.medschool.umaryland.edu/profiles/jiang-feng/

### Author Declarations

The study received approval from the Institutional Review Board of the University of Maryland Baltimore.

## REFERENCES

1. Mithoowani H, Febbraro M: Non-Small-Cell Lung Cancer in 2022: A Review for General Practitioners in Oncology, Curr Oncol 2022, 29:1828–1839

2. Schabath MB, Cress D, Munoz-Antonia T: Racial and Ethnic Differences in the Epidemiology and Genomics of Lung Cancer, Cancer Control 2016, 23:338–346

3. Ryan BM: Lung cancer health disparities, Carcinogenesis 2018, 39:741–751

4. Zanetti KA, Wang Z, Aldrich M, Amos CI, Blot WJ, Bowman ED, Burdette L, Cai Q, Caporaso N, Chung CC, Gillanders EM, Haiman CA, Hansen HM, Henderson BE, Kolonel LN, Marchand LL, Li S, McNeill LH, Ryan BM, Schwartz AG, Sison JD, Spitz MR, Tucker M, Wenzlaff AS, Wiencke JK, Wilkens L, Wrensch MR, Wu X, Zheng W, Zhou W, Christiani D, Palmer JR, Penning TM, Rieber AG, Rosenberg L, Ruiz-Narvaez EA, Su L, Vachani A, Wei Y, Whitehead AS, Chanock SJ, Harris CC: Genome-wide association study confirms lung cancer susceptibility loci on chromosomes 5p15 and 15q25 in an African-American population, Lung Cancer 2016, 98:33–42

5. Lerner L, Winn R, Hulbert A: Lung cancer early detection and health disparities: the intersection of epigenetics and ethnicity, J Thorac Dis 2018, 10:2498–2507

6. Cheng H, Hosgood HD, Deng L, Ye K, Su C, Sharma J, Yang Y, Halmos B, Perez-Soler R: Survival Disparities in Black Patients With EGFR-mutated Non-small-cell Lung Cancer, Clin Lung Cancer 2020, 21:177–185

7. Zingone A, Sinha S, Ante M, Nguyen C, Daujotyte D, Bowman ED, Sinha N, Mitchell KA, Chen Q, Yan C, Loher P, Meerzaman D, Ruppin E, Ryan BM: A comprehensive map of alternative polyadenylation in African American and European American lung cancer patients, Nat Commun 2021, 12:5605

8. Pine SR, Mechanic LE, Enewold L, Bowman ED, Ryan BM, Cote ML, Wenzlaff AS, Loffredo CA, Olivo-Marston S, Chaturvedi A, Caporaso NE, Schwartz AG, Harris CC: Differential Serum Cytokine Levels and Risk of Lung Cancer Between African and European Americans, Cancer Epidemiol Biomarkers Prev 2016, 25:488–497

9. Zavala VA, Bracci PM, Carethers JM, Carvajal-Carmona L, Coggins NB, Cruz-Correa MR, Davis M, de Smith AJ, Dutil J, Figueiredo JC, Fox R, Graves KD, Gomez SL, Llera A, Neuhausen SL, Newman L, Nguyen T, Palmer JR, Palmer NR, Perez-Stable EJ, Piawah S, Rodriquez EJ, Sanabria-Salas MC, Schmit SL, Serrano-Gomez SJ, Stern MC, Weitzel J, Yang JJ, Zabaleta J, Ziv E, Fejerman L: Cancer health disparities in racial/ethnic minorities in the United States, Br J Cancer 2021, 124:315–332

10. Santosh B, Varshney A, Yadava PK: Non-coding RNAs: biological functions and applications, Cell Biochem Funct 2015, 33:14–22

11. Distefano R, Nigita G, Le P, Romano G, Acunzo M, Nana-Sinkam P: Disparities in Lung Cancer: miRNA Isoform Characterization in Lung Adenocarcinoma, Cancers (Basel) 2022, 14:

12. Lin Y, Holden V, Dhilipkannah P, Deepak J, Todd NW, Jiang F: A Non-Coding RNA Landscape of Bronchial Epitheliums of Lung Cancer Patients, Biomedicines 2020, 8:

13. Su Y, Guarnera MA, Fang H, Jiang F: Small non-coding RNA biomarkers in sputum for lung cancer diagnosis, Mol Cancer 2016, 15:36

14. Gao L, Ma J, Mannoor K, Guarnera MA, Shetty A, Zhan M, Xing L, Stass SA, Jiang F: Genome-wide small nucleolar RNA expression analysis of lung cancer by next-generation deep sequencing, Int J Cancer 2015, 136:E623–629

15. Liao J, Yu L, Mei Y, Guarnera M, Shen J, Li R, Liu Z, Jiang F: Small nucleolar RNA signatures as biomarkers for non-small-cell lung cancer, Mol Cancer 2010, 9:198

16. Li N, Ma J, Guarnera MA, Fang H, Cai L, Jiang F: Digital PCR quantification of miRNAs in sputum for diagnosis of lung cancer, J Cancer Res Clin Oncol 2014, 140:145–150

17. Su Y, Fang H, Jiang F: Integrating DNA methylation and microRNA biomarkers in sputum for lung cancer detection, Clin Epigenetics 2016, 8:109

18. Su J, Liao J, Gao L, Shen J, Guarnera MA, Zhan M, Fang H, Stass SA, Jiang F: Analysis of small nucleolar RNAs in sputum for lung cancer diagnosis, Oncotarget 2016, 7:5131–5142

19. Xing L, Su J, Guarnera MA, Zhang H, Cai L, Zhou R, Stass SA, Jiang F: Sputum microRNA biomarkers for identifying lung cancer in indeterminate solitary pulmonary nodules, Clin Cancer Res 2015, 21:484–489

20. Shen J, Liao J, Guarnera MA, Fang H, Cai L, Stass SA, Jiang F: Analysis of MicroRNAs in sputum to improve computed tomography for lung cancer diagnosis, J Thorac Oncol 2014, 9:33–40

21. Xie Y, Todd NW, Liu Z, Zhan M, Fang H, Peng H, Alattar M, Deepak J, Stass SA, Jiang F: Altered miRNA expression in sputum for diagnosis of non-small cell lung cancer, Lung Cancer 2010, 67:170–176

22. Geng X, Tsou JH, Stass SA, Jiang F: Utilizing MiSeq Sequencing to Detect Circulating microRNAs in Plasma for Improved Lung Cancer Diagnosis, Int J Mol Sci 2023, 24:

23. Leng Q, Wang Y, Jiang F: A Direct Plasma miRNA Assay for Early Detection and Histological Classification of Lung Cancer, Transl Oncol 2018, 11:883–889

24. Leng Q, Lin Y, Jiang F, Lee CJ, Zhan M, Fang H, Wang Y: A plasma miRNA signature for lung cancer early detection, Oncotarget 2017, 8:111902–111911

25. Shen J, Liu Z, Todd NW, Zhang H, Liao J, Yu L, Guarnera MA, Li R, Cai L, Zhan M, Jiang F: Diagnosis of lung cancer in individuals with solitary pulmonary nodules by plasma microRNA biomarkers, BMC Cancer 2011, 11:374

26. Shen J, Todd NW, Zhang H, Yu L, Lingxiao X, Mei Y, Guarnera M, Liao J, Chou A, Lu CL, Jiang Z, Fang H, Katz RL, Jiang F: Plasma microRNAs as potential biomarkers for non-small-cell lung cancer, Lab Invest 2011, 91:579–587

27. Shen J, Stass SA, Jiang F: MicroRNAs as potential biomarkers in human solid tumors, Cancer Lett 2013, 329:125–136

28. Shen J, Jiang F: Applications of MicroRNAs in the Diagnosis and Prognosis of Lung Cancer, Expert Opin Med Diagn 2012, 6:197–207

29. Ma J, Li N, Lin Y, Gupta C, Jiang F: Circulating Neutrophil MicroRNAs as Biomarkers for the Detection of Lung Cancer, Biomark Cancer 2016, 8:1–7

30. Lin Y, Leng Q, Zhan M, Jiang F: A Plasma Long Noncoding RNA Signature for Early Detection of Lung Cancer, Transl Oncol 2018, 11:1225–1231

31. Gupta C, Su J, Zhan M, Stass SA, Jiang F: Sputum long non-coding RNA biomarkers for diagnosis of lung cancer, Cancer Biomark 2019, 26:219–227

32. Li N, Dhilipkannah P, Jiang F: High-Throughput Detection of Multiple miRNAs and Methylated DNA by Droplet Digital PCR, J Pers Med 2021, 11:

33. Ma J, Li N, Guarnera M, Jiang F: Quantification of Plasma miRNAs by Digital PCR for Cancer Diagnosis, Biomark Insights 2013, 8:127–136

34. Esfandi F, Taheri M, Omrani MD, Shadmehr MB, Arsang-Jang S, Shams R, Ghafouri-Fard S: Expression of long non-coding RNAs (lncRNAs) has been dysregulated in non-small cell lung cancer tissues, BMC Cancer 2019, 19:222

35. Boeri M, Verri C, Conte D, Roz L, Modena P, Facchinetti F, Calabro E, Croce CM, Pastorino U, Sozzi G: MicroRNA signatures in tissues and plasma predict development and prognosis of computed tomography detected lung cancer, Proc Natl Acad Sci U S A 2011, 108:3713–3718

36. Anjuman N, Li N, Guarnera M, Stass SA, Jiang F: Evaluation of lung flute in sputum samples for molecular analysis of lung cancer, Clin Transl Med 2013, 2:15

37. Yu L, Todd NW, Xing L, Xie Y, Zhang H, Liu Z, Fang H, Zhang J, Katz RL, Jiang F: Early detection of lung adenocarcinoma in sputum by a panel of microRNA markers, Int J Cancer 2010, 127:2870–2878

38. Aberle DR, Adams AM, Berg CD, Black WC, Clapp JD, Fagerstrom RM, Gareen IF, Gatsonis C, Marcus PM, Sicks JD: Reduced lung-cancer mortality with low-dose computed tomographic screening, N Engl J Med 2011, 365:395–409

39. Mitchell KA, Zingone A, Toulabi L, Boeckelman J, Ryan BM: Comparative Transcriptome Profiling Reveals Coding and Noncoding RNA Differences in NSCLC from African Americans and European Americans, Clin Cancer Res 2017, 23:7412–7425

40. Wang Y, Shang S, Yu K, Sun H, Ma W, Zhao W: miR-224, miR-147b and miR-31 associated with lymph node metastasis and prognosis for lung adenocarcinoma by regulating PRPF4B, WDR82 or NR3C2, PeerJ 2020, 8:e9704

41. Rivera MP, Gudina AT, Cartujano-Barrera F, Cupertino P: Disparities Across the Continuum of Lung Cancer Care, Clin Chest Med 2023, 44:531–542

42. Harrison S, Judd J, Chin S, Ragin C: Disparities in Lung Cancer Treatment, Curr Oncol Rep 2022, 24:241–248

43. Haddad DN, Sandler KL, Henderson LM, Rivera MP, Aldrich MC: Disparities in Lung Cancer Screening: A Review, Ann Am Thorac Soc 2020, 17:399–405

44. Yang D, Wang JJ, Li JS, Xu QY: miR-103 Functions as a Tumor Suppressor by Directly Targeting Programmed Cell Death 10 in NSCLC, Oncol Res 2018, 26:519–528

45. Di Paolo D, Pontis F, Moro M, Centonze G, Bertolini G, Milione M, Mensah M, Segale M, Petraroia I, Borzi C, Suatoni P, Brignole C, Perri P, Ponzoni M, Pastorino U, Sozzi G, Fortunato O: Cotargeting of miR-126-3p and miR-221-3p inhibits PIK3R2 and PTEN, reducing lung cancer growth and metastasis by blocking AKT and CXCR4 signalling, Mol Oncol 2021, 15:2969–2988

46. Lebanony D, Benjamin H, Gilad S, Ezagouri M, Dov A, Ashkenazi K, Gefen N, Izraeli S, Rechavi G, Pass H, Nonaka D, Li J, Spector Y, Rosenfeld N, Chajut A, Cohen D, Aharonov R, Mansukhani M: Diagnostic assay based on hsa-miR-205 expression distinguishes squamous from nonsquamous non-small-cell lung carcinoma, J Clin Oncol 2009, 27:2030–2037

47. Pirlog R, Chiroi P, Rusu I, Jurj AM, Budisan L, Pop-Bica C, Braicu C, Crisan D, Sabourin JC, Berindan-Neagoe I: Cellular and Molecular Profiling of Tumor Microenvironment and Early-Stage Lung Cancer, Int J Mol Sci 2022, 23:

48. Zhao YL, Zhang JX, Yang JJ, Wei YB, Peng JF, Fu CJ, Huang MH, Wang R, Wang PY, Sun GB, Xie SY: MiR-205-5p promotes lung cancer progression and is valuable for the diagnosis of lung cancer, Thorac Cancer 2022, 13:832–843

